# Tucaresol: A Clinical Stage Oral Candidate Drug With Two Distinct Antiviral Mechanisms

**DOI:** 10.1101/2024.09.14.24312736

**Authors:** Christopher L. Penney, Boulos Zacharie, Jean-Simon Duceppe

## Abstract

Globally, approximately 39 million people are living with Human Immunodeficiency Virus, HIV, arising from approximately 86 million infections since this epidemic began in 1981. However, the number of HIV infections is unevenly distributed with two thirds of global infections confined to Sub-Saharan Africa. Due to viral drug resistance, the most effective treatment requires a triple drug combination thereby adding to the complexity and cost of therapy. As such, many people living with HIV or at risk of infection do not have access to prevention or treatment of this potentially fatal disease. There is no cure for HIV [1]. Tucaresol is an orally active clinical stage drug which functions as a host targeted antiviral agent by protection or reconstitution of CD4+ T helper immune cells. We report herein that Tucaresol also displays in-vitro activity against HIV in infected human peripheral blood mononuclear cells. Although this in-vitro antiviral activity is not potent, the excellent safety profile and bioavailability of Tucaresol, along with its low Molecular Weight, support attainment of relevant drug concentrations in man to achieve significant in-vivo activity. This is demonstrated by previously reported stabilization of viremia in a prior proof of concept phase 1b/2a HIV clinical trial [2]. It is possible that the significant in-vivo activity of Tucaresol arises from synergy between co-stimulation of CD4+ T helper cells and the direct activity against virally infected cells. A pan in-vitro viral screen of Tucaresol further revealed a weak, direct antiviral activity against human herpes virus 6B, human papillomavirus 11, measles virus and hepatitis B virus.

## Background

In 1987 Azidothymidine, AZT, was the first drug approved by the US FDA for treating AIDS. Since then, multiple drugs targeting various viral targets have been approved for use. Anti-HIV drug classes include Nucleoside Reverse Transcriptase Inhibitors, Non-Nucleoside Reverse Transcriptase Inhibitors, Protease Inhibitors, Fusion Inhibitors, CCR5 Antagonists, Integrase Inhibitors, Attachment/Post Attachment Inhibitors, Capsid Inhibitors, PK Enhancers and combination HIV medicines containing two or more HIV drugs from one or more drug classes. Subsequently, the FDA has approved individual and combination drug therapies representing approximately 50 different treatment modalities against HIV as listed in [3]. In spite of a broad selection of approved HIV drugs against multiple viral targets and very few host-targets, there is no cure for HIV. An important issue pertaining to disease pathology is that the HIV virus rapidly mutates. This requires the presence of two or more antiviral drugs with distinctly different mechanisms of antiviral activity in order to have a significant impact upon viral infections. Thus, for example, treatment with AZT alone could lead to drug resistance within a few days while treatment with a combination of AZT and dideoxycytidine, ddC, demonstrated that this drug regimen was more effective than AZT alone in preventing decline in CD4+ T helper cells and death. Therefore, while the efficacy of two drug combination therapy was superior to monotherapy, the improved drug efficacy was of limited duration. However, the discovery of additional drugs, notably HIV protease inhibitors, facilitated triple drug combination therapy or Highly Active Antiretroviral Therapy, HAART [4]. Triple drug combination therapy can durably suppress viral replication to minimal levels while further retarding the development of drug resistance. In order to offer the most efficacious, safest triple drug combination therapy, attention must be given to the fact that the success achieved thus far regarding treatment of HIV, without a permanent cure of the disease, is at risk of being undermined by development of antiviral drug resistance. Therefore, there is a need for HIV drugs that target the viral life cycle at stages not routinely targeted by approved HIV drugs. Thus, for example, a candidate drug is reported [5] that apparently is a first-in-class inhibitor of HIV virus production by targeting a virus - host (2 host enzymes) complex important for HIV - Gag assembly which facilitates viral capsid assembly. Additionally, a forceful assault on HIV requires a combination drug regimen wherein the druggable or biochemical target is as distinct as possible from the other member(s) of the combination therapy in order to suppress drug resistance as much as possible. Particularly distinct is a drug combination where one drug targets the virus while the other drug targets a host protein shared with the virus. Therapy becomes ineffective upon appropriate mutation of the HIV virus because said mutation potentially permits full function of the virus if the virus no longer needs the mutated biochemical target. However, the virus is trapped since it cannot mutate to avoid or dampen the host targeted protein since the virus is not genetically coded to inactivate or circumvent the host target protein. In fact, only one FDA approved drug is available for use in combination therapy. The HIV host targeted drug Maraviroc targets the entry coreceptor CCR5 [6]. Alternatively, the most effective antiviral drug may be a candidate drug wherein the two distinct biochemical target, the viral target and the host target, are present within the same molecule. The work reported herein suggests that this is the case with the HIV clinical stage candidate drug, Tucaresol.

### Tucaresol; A Clinical Stage Candidate Drug For Treatment Of HIV

Tucaresol (Figure 1), or 4-[(2-formyl-3-hydroxyphenoxy) methyl] benzoic acid (IUPAC nomenclature; Molecular Weight = 272.25 g/mol) is an orally active antiviral drug with good oral bioavailability (70%), ambient temperature stability and excellent potency at a daily dose of less than 100 mg. In addition to the favorable potency of Tucaresol, ample product is readily produced by a one-step synthesis developed and later modified by us [7]. Importantly, Tucaresol is not encumbered by pre-existing patents claiming compound ownership (Composition of Matter), synthesis (process) or treatment of HIV.

**Figure 1.**
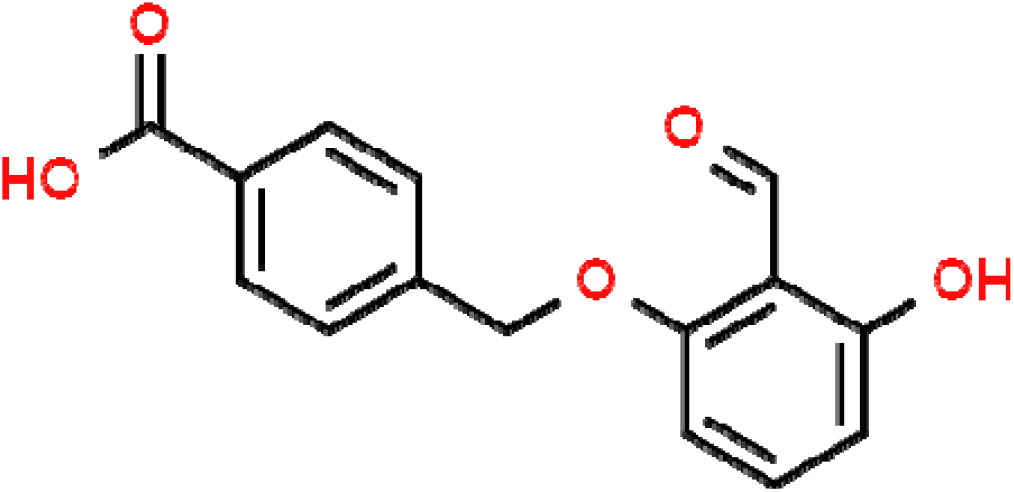
Structure of Tucaresol (free acid)

Tucaresol was under development years ago by Glaxo Wellcome for the palliation of sickle cell anemia. Since the drug mechanism required stoichiometric binding of Tucaresol to hemoglobin to prevent red blood cell sickling and increase oxygen affinity, Tucaresol was administered to patients in high doses. Subsequently, human toxicity and pharmacokinetic data, such as maximum tolerated dose, were available well above that required for clinical work with Tucaresol as an antiviral agent at daily doses of up to 100 mg per patient. However, it was during this work with high dose Tucaresol that indications of immune activity was observed, thereby resulting in cessation of the sickle cell anemia project. Exploration of this newly discovered immunological activity revealed that Tucaresol functions as a host targeted antiviral agent by protection of antiviral immune cells via controlled co-stimulation of specific T-cells (CD4+ T helper cells) in the presence of HIV or other pathogenic virus to obtain normal immune status or by reconstitution of the same antiviral immune cells significantly depleted by a pathogenic virus, as has been demonstrated in HIV patients (see below) and SIV infected macaques. Indeed, the process of T-cell dysfunction, T-cell exhaustion, represents an important pathway by which the pathogenic virus subverts T-cell function. Important to note is that Tucaresol does not function as an immunostimulant in normal immune status individuals thereby preventing a hyperactive and potentially fatal response such as cytokine storm (cytokine release syndrome). In fact, the ability to stimulate a controlled immune response in an immune deficient mammal such as may occur during a viral infection is a key tenant of the intellectual property of Tucaresol as defined by the granted patents to Glaxo Wellcome Inc. These patents issued in 1996, 1999 and 2000 respectively state in their first claim a method for treating a immunodeficient mammal, which exemplifies the use of Tucaresol to treat and eliminate (in one macaque) an SIV infection; US patents 5.508,310, 5,872,151 and 6,096,786.

### Tucaresol; HIV Clinical Studies

In view of the important role played by T-cells regarding patient survivability during an HIV infection and with regard to binding of Tucaresol to CD4+ T helper target cells, after completion of requisite PK/PD and animal safety studies, a phase 1 clinical safety trial was undertaken. This was followed by initiation of a 45 HIV patient phase 2 trial with patients already receiving HAART (Highly Active Antiviral Therapy) and Tucaresol at a daily dose of 25 mg or 50 mg for 3 months. This study was not completed and no results reported [8]. However, significant modulation of T-cell activity by Tucaresol in an HIV phase 1b/2a trial was published in 2004 as a GlaxoSmithKline and University of Milan collaboration [2]. This clinical trial consisted of 4 groups of HIV positive patients, a total of 24 patients, in which half of the patients received HAART prior to the trial and the other half of the patients were HAART naive. This was a 16 week pulse dose escalation protocol in which Tucaresol was administered as one 25 mg dose during week 1, 25 mg/day for 4 days during week 4, 50 mg/day for 4 days during week 8 and 100 mg/day for 4 days during week 12 with time between doses to permit drug wash-out. One of the 4 groups received only Tucaresol while the second group received Tucaresol and HAART at the same time. Groups 3 and 4, already on HAART prior to the trial, received Tucaresol according to the dose schedule above. The difference between the two groups is that one group consists of patients that are immunological nonresponders as evidenced by a below average CD4+ T-helper cell count. Following administration of Tucaresol,increases in percentages of memory T lymphocytes (CD4+/CD45R0+) was observed in all patients, including those patients treated only with Tucaresol. Additionally, a significant increase in memory T-cells on week 12 in the group already on HAART and on week 13 in the group of immunological non-responders was observed, in both cases p < 0.05. Any significant p value is of note since each group consists of only 6 patients. More significant was a sustained increase in percentages of naive T lymphocytes (CD4+/CD45RA+) observed in all patients concomitantly with administration of Tucaresol. Also, an increase in naive CD4+ T-cells was observed in approximately week 8 in all groups following the third Tucaresol administration. Env-stimulated perforin-expressing CD8+ cytotoxic T-cells were increased in all groups. Perforin-expressing p24-stimulated cytotoxic CD8+ T-cells were similarly amplified. As regards HIV plasma viremia, there were no changes in HIV RNA levels, including patients treated only with Tucaresol, except a decrease in HIV RNA in starting simultaneously treatment with Tucaresol and HAART. Therefore, while treatment with Tucaresol did not eliminate viremia, it prevented a significant increase in HIV viremia. Administration of Tucaresol was associated with increased interleukin 12 and decreased interleukin 10. In particular, the reduction of interleukin 10 upon administration of Tucaresol was highly significant at weeks 8, 12 and 16 for patients already on HAART; p<0.001. Interleukin 10 reversibly inhibits virus-specific T-cells and high interleukin 10 expression can predict poor clinical outcomes in HIV patients [9]. No severe adverse effects (SAEs) were observed in patients administered Tucaresol while under treatment with HAART therapy. Otherwise, the only observed SAE was in two viremic patients who experienced lymphadenopathy. Approximately a decade after publication of the Tucaresol phase 1b/2a HIV clinical results, a 2015 report announced “Dawn of antioxidants and immune modulators to stop HIV-progression and boost the immune system in HIV/AIDS patients” [10]. It was noted that HIV patients may experience oxidative stress which can lead to exacerbation of HIV, enhancement of HIV replication or augmentation of apoptosis. Interestingly, the core structure of Tucaresol consists of a dihydroxybenzaldehyde scaffold which possesses antioxidant activity. For comparison, it is published that 3-hydroxybenzaldehyde is a more potent antioxidant than vitamin C [11]. Similarly, it has been noted that some HIV patients on HAART therapy do not readily reconstitute their immunity, as demonstrated by a low CD4+ T-cell count, despite their full virologic response to HAART. This impaired immune response is linked to increased risk of disease progression and death [12] thereby adding importance to the addition of Tucaresol to the HAART HIV treatment regimen. In conclusion, in patients on HAART with proper viral suppression, treatment with Tucaresol resulted in (1) stimulation of cytotoxic T-lymphocyte activity (2) generation of naive T-cells and (3) did not result in any adverse effects or increase in patient viral load.

## Results

### Direct Antiviral (Non-Immunologic) Activity Of Tucaresol

Tucaresol functions as a host targeted antiviral agent by co-stimulation of CD4+ T helper cells which eliminate pathogenic viruses by mounting a sufficient, protective immune response without excessive, dangerous hyperactivation of the immune system. For more than 30 years multiple papers have been published regarding the immunological properties of Tucaresol as an antiviral agent, anticancer agent and vaccine adjuvant. The well described immunomodulatory properties and safety profile of Tucaresol prompted us to propose combination of Tucaresol with approved or candidate drugs for treatment of covid-19 (SARS-CoV-2) virus, per our published review article [13]. However, the results published on the phase 1b/2a HIV clinical trial of Tucaresol [2] demonstrated Tucaresol has a significant effect on T-cell function and numbers in humans as indicated by appropriate T-cell and interleukins 2, 10 and 12 assays. For example, with a small group of patients already on HAART therapy, n = 6, “IL-10 was considerably reduced” at weeks 8, 12 and 16 after the start of treatment with Tucaresol, p<0.001. This is important since interleukin 10 correlates positively with viral load and diminishes after successful antiretroviral therapy [9]. Additionally, during chronic viral infection it has been reported that interleukin 10 directly inhibits CD8+ T cell function by enhancing N-glycan branching on T-cell surface glycoproteins thereby by decreasing antigen sensitivity or T-cell binding to the virus [14]. Also, at the end of this 16 week clinical trial there was no change in patients HIV RNA levels except a decrease in HIV RNA in the group of patients starting simultaneous treatment with Tucaresol and HAART. That is, Tucaresol was able to stabilize or control HIV viremia. In view of these positive results obtained with a small group of patients, especially the significant decrease in interleukin 10 and maintenance of a constant level of viremia, the question arises if these results reflect two simultaneous antiviral mechanisms inherent to Tucaresol? Two antiviral mechanisms which are the well documented co-stimulation of CD4+ immune cells and a direct antiviral mechanism against HIV? The direct antiviral activity of Tucaresol appears weak, as determined by the effective concentration of Tucaresol required to inhibit HIV in infected human peripheral blood mononuclear cells (PBMCs) in 7 day cell culture; EC50 = 35 *µ*M. However, this in-vitro result does not preclude the possibility that synergy exists in-vivo between the two antiviral mechanisms thereby resulting in significant clinical efficacy, including stabilized HIV viremia by Tucaresol. Further evidence for a direct, non-immunologic mechanism of antiviral activity is the observation that there is no increase in viremia in the group of immune non-responder patients, n = 6, with below normal CD4+ immune cell count. Indeed, a few hydroxybenzaldehyde compounds are reported to possess direct in-vitro antiviral activity. 2-Hydroxybenzaldehyde (salicylaldehyde) and 2,3-dihydroxybenzaldehyde were observed to suppress replication of herpes simplex virus (HSV-1) in cell culture with salicylaldehyde, IC50 = 22 *µ*M [15]. Recently, 4-hydroxy-3-methoxybenzaldehyde (vanillin) was reported to inhibit measles virus in cell culture with IC50 = 5 *µ*M [16].

Therefore, Tucaresol’s 2,6-dihydroxybenzaldehyde scaffold is likely the key portion of the molecule responsible for the direct antiviral activity of Tucaresol. Furthermore, this in-vitro antiviral activity in HIV infected PBMCs likely cannot be attributed solely to the antioxidant properties of the 2,6-dihydroxybenzaldehyde scaffold since a pan in-vitro viral screen of Tucaresol in 14 viral cultures revealed that only 5 of the viruses were susceptible to the direct in-vitro antiviral activity of Tucaresol with EC values ranging from 7.5 - 37 *µ*M. This in-vitro evaluation of Tucaresol against 14 viruses is summarized below with EC50 (Effective Concentration) data for each virus. The cut-off value for a quantifiable EC50 was EC50 < 50 *µ*M since any value significantly greater than 50 *µ*M was viewed as not meaningful for a potential therapeutic agent. With an EC50 = 35 *µ*M and assuming a blood volume of 5 liters with Tucaresol exclusively targeting the lymphatics (not strictly correct) the amount of Tucaresol required for each oral dose to attain EC50 is manageable at approximately 50 mg. Support for the in-vitro evaluation of Tucaresol against the 14 viruses listed below was provided by the US National Institute of Health, NIH/NIAID, with testing undertaken at labs contracted by NIH. A summary of this study follows.

#### 1) In-vitro Evaluation of Tucaresol; Human Immunodeficiency Virus

Human subjects were not recruited for any part of this study. Human PBMCs were isolated from whole blood buffy coats purchased from BioIVT. Tucaresol was prepared as a 10 mM soluble DMSO stock solution. The clinical strain of HIV-1BR/92/004 **was** obtained from the AIDS Research and Reference Reagent Program, Rockville, Maryland. Two fold concentration of Tucaresol in assay media was added in triplicate to a predetermined dilution of virus. The whole was incubated for 30 minutes at 37°C/5% CO2. Following this incubation, appropriately prepared PBMCs that were induced to proliferate with PHA-P (Phytohemaglutinin) were resuspended in fresh tissue culture medium and plated in a 96 well microtiter plate (50 microliters/well). After 7 days in culture at 37°C/5% CO2. HIV replication was quantified by the measurement of cell free HIV reverse transcriptase activity in the tissue culture supernatant. Cytotoxicity was evaluated with the tetrazolium dye XTT. By means of these in-vitro assays, the EC50 value and the CC50 value for Tucaresol was determined as 35 and >50 *µ*M respectively yielding a Therapeutic Index >1.4. AZT was evaluated in parallel as a positive control (HIV inhibitor) and performed as expected with EC50 = 0.42 nM. In spite of the multiple publications regarding Tucaresol over the years, to the best of our knowledge there is no public disclosure regarding the direct antiviral (non-immunologic) activity of Tucaresol as ascertained by in-vitro assay of Tucaresol with HIV infected human PBMCs. Multiple publications have appeared regarding the immunologic properties and mechanism of action of Tucaresol. This included the first in-vitro assessment of the immunological activity of Tucaresol in the presence of antigen and mitogen-stimulated proliferation and cytokine production by PBMCs of HIV-infected individuals and healthy controls [17]. Based on these results supportive of a pro-inflammatory Th1 immune response, it was suggested Tucaresol would be particularly useful for treatment of viral diseases characterized by defective CMI (cell mediated immunity) such as HIV and hepatitis B. Important was the fact that Tucaresol-induced immune activation has no effect on viral replication [17]. The sponsor for this study was RTI International with testing undertaken at ImQuest BioSciences Inc. Frederick, Maryland.

#### 2) In-vitro Evaluation of Tucaresol; Hepatitis B Virus

A microtiter plate assay format using a liver derived AD38 cell line that inducibly expresses the hepatitis B genome was used to determine the antiviral activity of Tucaresol against hepatitis B. This assay format consisted of a primary screen in which antiviral activity was expressed as visible cytopathic effect (CPE) induced by the virus on the AD38 liver cells followed by determination of the reduced CPE in the presence of Tucaresol, expressed as EC50. Depending upon the cytopathic effect observed in the presence of Tucaresol, a secondary hepatitis B assay was performed to determine the release of extracellular hepatitis B viral DNA with qPCR (quantitative polymerase chain reaction) and the release of viral hepatitis B surface antigen with peroxidase conjugated anti-hepatitis B surface antigen monoclonal antibody. The first primary antiviral assay result indicated that Tucaresol has direct antiviral activity against hepatitis B; EC50 < 0.32 *µ*M. The first secondary hepatitis B surface antigen (HBsAg) assay indicated a reduction in hepatitis B virus with a reduced concentration of HBsAg virus particles; EC50 (IC50) = 1.9 *µ*M. In order to determine a precise EC50 value for the primary assay, it was repeated three times to yield the same result; EC50 > 5 *µ*M. Similar values were obtained for the three repeats of the release of HBsAg into the culture media; EC50 = 7 *µ*M and twice EC50 > 5 *µ*M. Interestingly, examination of the raw assay data revealed that instead of a reduction in viral DNA release into the culture media, the increase of viral DNA release relative to the beginning of the culture (time zero) was a 5.2 fold increase of viral DNA. As noted above, the accompanying viral HBsAg data did not indicate a significant increase in release of HBsAg virus particles into the culture media. This behavior of limited or no reduction of HBsAg viral particles has been reported for a specific class of direct (non-immunologic) antiviral drugs; CAMs or Capsid Assembly Modulators (Inhibitors). For example, in the chronic hepatitis B phase 2 clinical trial of the J & J (Janssen) CAM candidate hepatitis B drug Bersacapavir, it was reported that there was limited reduction or no effect on release of HBsAg virus particles [18]. This assay result is in agreement with the four HBsAg results noted above for Tucaresol, two minor reductions of HBsAg; EC50 = 1.9 and 7 *µ*M and two assay results indicating no observable reduction in HBsAg. However, it was also reported that there was a pronounced reduction of extracellular viral DNA in the presence of Bersacapavir [18] which is clearly contrary to the significant increase in viral DNA observed in the assay media in the presence of Tucaresol. A possible explanation for the similar HBsAg results but contrary viral DNA results obtained in the in-vitro assay of Tucaresol versus the results reported for Bersacapavir is that the viral DNA release into the assay media occurs later in the presence of Tucaresol relative to when it takes place in the presence of Bersacapavir. That is, a more fully completed viral DNA molecule is released into the assay media that is detectable in the antiviral assay of Tucaresol but the less completed viral DNA molecule is not detectable in the antiviral assay of Bersacapavir. A fifth, final Tucaresol primary assay was undertaken with a fresh sample of Tucaresol to eliminate the possibility that degradation of Tucaresol had not taken place in the stock solution. Once again, an EC50 > 5 *µ*M was obtained for the release of viral DNA into the assay media. Extension of the concentration range for the assay of viral DNA revealed EC50 > 32 *µ*M. However, the increase of viral DNA released into the culture media was similar to that observed for prior assays; 4.5 fold increase of viral DNA relative to the concentration at time zero. Immunosuppressive anticancer agents such as doxorubicin are known to cause an increase in cell culture viral DNA but this is accompanied by an increase in cell culture HbsAg. The sponsor for this study was University of Southern Utah with testing undertaken at ImQuest BioSciences Inc. Frederick, Maryland. Virus Strain, HepAD38. Cell Line, AD38.

#### 3) In-vitro Evaluation of Tucaresol; Human Herpes 6B

Roseola is a common, contagious viral infection caused by human herpesvirus 6. Interestingly, Tucaresol was observed to be slightly more potent, EC50 = 7.5 *µ*M, than the assay standard Cidofovir; EC50 = 10.3 *µ*M. This suggests investigation of Tucaresol as an oral and/or topical treatment for roseola, including roseola associated rash. Testing was at University of Alabama. Assay, Quantitative polymerase chaIn reaction. Virus Strain, Z2g.Cell Line, MOLT-3.

#### 4) In-vitro Evaluation of Tucaresol; Human Papillomavirus 11

With more than 9 out of 10 cervical cancer cases caused by human papilloma virus (HPV), most cervical cancer can be prevented by use of the papilloma vaccine. One of the few topical treatments for HPV is the immunostimulant Imiquimod but it has a comparable rating with the classic anticancer drug, topical 5-Fluorouracil. Tucaresol was approximately two fold less potent against HPV than Herpes virus 6B; EC50 = 18.4 *µ*M versus EC50 = 7.5 *µ*M. For comparison with the HPV assay standard, Tucaresol was EC50 = 18.4 *µ*M while the positive guanine assay standard was EC50 = 2.7 *µ*M. This suggests investigation of Tucaresol as an oral and/or topical treatment for cervical cancer. Testing was at University of Alabama. Assay, Nano-Glo Luciferase. Virus Strain, HE611260.1. Cell Line, C-33A.

#### 5) In-vitro Evaluation of Tucaresol; Measles Virus (MeV)

Measles is a highly contagious respiratory virus for which there is no treatment. However, it is mostly preventable with appropriate vaccination. The in-vitro efficacy of Tucaresol against measles virus was similar to the efficacy determined against HIV; EC50 = 37 *µ*M versus EC50 = 35 *µ*M. A cellular immune response is important for induction of protective immunity against measles since this virus can suppress cell mediated immunity thereby increasing viremia. This suggests investigation of Tucaresol as a potential immunomodulator to facilitate rapid recovery from infection and/or induction of virus induced immune amnesia with accompanying secondary infection. Testing was at University of Southern Utah. Assay, Viral induced visible cytopathic effect (CPE). Virus Strain, CC. Cell Line, Vero 76.

#### 6) In-vitro Evaluation of Tucaresol: Viruses Without Observable In-vitro Activity

A pan viral screen of Tucaresol revealed a weak, direct in-vitro antiviral activity, EC50 < 50 *µ*M, against 5 infectious viruses. These viruses are 1. HIV, 2. Human Herpesvirus 6,3. Human Papillomavirus 11, 4. Measles, 5. Hepatitis B. However, evaluation of 9 other infectious viruses did not identify quantifiable activity. These viruses are 1. Influenza A, H1N1 (swine flu), 2. Rift Valley Fever Virus, 3. Adenovirus 5, 4. Varicella-Zoster Virus (chickenpox, later shingles). 5. Epstein-Barr Virus. 6. Cowpox Virus. 7. Vaccinia Virus. Herpes Simplex Virus 1. 9. Herpes Simplex Virus 2. The genetic material of the 5 viruses susceptible to direct antiviral activity of Tucaresol were RNA viruses (HIV, measles) or DNA viruses (hepatitis B, human herpes virus 6B, and human papilloma 11).

## Discussion

The clinical stage antiviral immunomodulator Tucaresol has been long studied as an antiviral agent, anticancer agent and a vaccine adjuvant. This selection of research areas was predicated on the unique ability of Tucaresol to adequately stimulate a suppressed immune system by sufficient stimulation of CD4+ T helper cells to return to normal immune status without a hyperactive immune response thereby preventing a dangerous response such as cytokine release syndrome (cytokine storm). However, none of the published literature indicated that Tucaresol has direct, non-immunologic, antiviral activity as a result of interaction with a virus such as HIV. The work described herein demonstrates that the in-vitro antiviral activity of Tucaresol interacting with select infectious viruses likely arises from a specific, weak binding interaction with viral proteins. A specific binding interaction between Tucaresol and a protein surface has been demonstrated regarding binding and subsequent protein conformational changes that resulted in the improved oxygen affinity of hemoglobin from patients with sickle cell anemia. It was published that the appropriate conformational change within the hemoglobin molecule arises from a noncovalent bridge linking Tucaresol with two hemoglobin alpha subunits via Schiff base formation with the terminal amino group of one alpha protein subunit and the aldehyde group of Tucaresol and an ionic bond (salt bridge) with the terminal amino group of the second alpha protein subunit and the carboxylate group of Tucaresol (19). A more relevant analogy to the binding and inactivation of viruses by Schiff base formation between a virus protein amino group and the aldehyde function of Tucaresol may be interaction of the aldehyde function(s) of formaldehyde or glutaraldehyde with viruses forming Schiff bases to function as a fixative (microscopy) or preparation of vaccines. A classic example of viral vaccines is the annual manufacture of influenza vaccine. Perhaps the ability of Tucaresol to inactivate certain viruses in-vitro is additionally or alternatively implicit to the 2,6-dihydroxybenzaldehyde portion (scaffold) present in Tucaresol. This is supported by the antiviral activity of vanillin against measles and salicylaldehyde and 2,3-dihydroxybenzaldehyde against herpes simplex 1. Finally, regarding the variable results obtained with the in-vitro assay of hepatitis B virus, it appears likely that Tucaresol exhibits in-vitro activity against hepatitis B. The variable HBsAg results obtained with four in-vitro assays in which two of the assays did exhibit a reduction in hepatitis B surface antigen, EC50 = 1.9 and 7 *µ*M, were consistent with direct antiviral drugs against hepatitis B that function as CAMs (Capsid Assembly Modulator), as noted above. However, the significant release of hepatitis B DNA into the media is not consistent with Tucaresol functioning as a CAM unless, as hypothesized above, the released hepatitis B DNA was close enough to completion to be detected by the hepatitis B DNA assay after release from the incomplete viral capsid into the media. Another explanation for the significant release of hepatitis B DNA into the media is that inthe case of hepatitis B virus, Tucaresol functions as an immunosuppressant thereby facilitating replication of the hepatitis B virus. However, if this is correct then the increase in released viral DNA should have been accompanied by a significant increase in HBsAg particles into the media, as noted to be the case with immunosuppressive anticancer drugs such as doxorubicin, which was clearly not the case with Tucaresol.. We therefore conclude that Tucaresol has a direct antiviral effect on hepatitis B virus, with a possible effect on viral capsid assembly. Interestingly, if it is correct that Tucaresol has two mechanisms of antiviral activity, immunologic as well as direct antiviral activity via inhibition of viral capsid assembly, than both mechanisms of antiviral activity could contribute to a cure for hepatitis B by their ability to inhibit viral cccDNA. Relatively recently, it was published that hepatitis B virus specific CD4+ T-cell responses differentiate a functional cure for hepatitis B from a chronic (surface antigen) hepatitis B infection [20]. This alone suggests that Tucaresol may be particularly useful either in combination therapy or as monotherapy to ensure an optimum T-cell immune response during treatment of hepatitis B Infection.

With demonstration herein of direct antiviral activity by Tucaresol against HIV in addition to herpes virus 6B, papilloma 11, measles and hepatitis B, in addition to the well-studied host targeted controlled immunostimulation of CD4+ T helper cells, Tucaresol may be a particularly useful candidate drug for treatment of these five viruses. Additionally, other factors such as excellent safety, good oral bioavailability, ease of manufacture and favorable pharmacoeconomics further contribute to the desirability of Tucaresol as an antiviral drug, especially in economically challenged situations. Finally, another opportunity exists for Tucaresol as the lead molecule for a series of analogs where Tucaresol itself functions as a core or for a series of new candidate drugs. Tucaresol would function as a scaffold to preserve the immune mechanism with its carboxyl group and/or hydroxyl group as points of attachment of functional groups to increase direct binding of Tucaresol to a target viral protein.

## Data Availability

All data produced in the present study are available upon reasonable request to the authors

## Declarations

No animal studies or human trials were performed by the authors for this project or by the US National Institute of Health or its’ contractors.

## Acknowledgement

We gratefully acknowledge the skillful guidance and participation of Amanda Ulloa, Virology Branch, NIAID/NIH. The literature contributions and “competitive intelligence” monitoring of Dr. Alan Cameron is greatly appreciated.

## Author Contributions

Project conceptualization CP, BZ, Drafting of original manuscript CP, Review and editing CP, BZ, JSD, Literature collection and analysis CP, BZ, Project administration CP,BZ, JSD, Methodology JSD, BZ, Experimentation JSD.

## Funding

The authors declare that no grants were received in support of this work.

## Conflicting Interest

The authors declare that there is no conflict of interest.

## Notes

### Competing Interest Statement

The authors have declared no competing interest.

### Funding Statement

This study did not receive any funding

### Author Declarations

Human subjects were not recruited for any part of this study. Human PBMCs were isolated from whole blood buffy coats purchased from BioIVT.

